# Bone Quality is Associated with Fragility Fracture in Patients with Hemoglobinopathies

**DOI:** 10.1101/2024.11.12.24317185

**Authors:** Fung Ellen B, Sarsour Iman, Manzo Raquel, Lal Ashutosh

## Abstract

**Background:** Low bone mass is common in adults with thalassemia (Thal) and sickle cell disease (SCD), though disease-specific artifacts may contribute to inaccuracies in bone mineral density (BMD) assessment. Trabecular bone score (TBS), an indicator of bone quality, is not susceptible to these challenges and may improve fracture risk prediction.

**Methods:** A retrospective chart review was conducted in patients with Thal or SCD who had at least one spine BMD scan by DXA performed in the past 10 years. The most recent scan was reanalyzed for bone quality using TBS Insight (Medimaps v 3.0.2) with abnormal defined as TBS <1.20. Fracture prevalence was determined by patient report with medical record validation. Patients were compared to healthy controls who participated in previous research.

**Results:** Data was abstracted from 126 patients with Thal (31.7±11.9 yrs, 51% Male), 170 with SCD (24.6±13.5 yrs, 43% Male), and 64 controls (25.9±8.0 yrs, 17% Male). Low bone mass was observed in 63% of Thal compared with 36% of SCD and 3% of controls (p<0.001); while only 15% of patients had abnormal TBS. History of fracture was present in 35.6% of Thal and 22.9% of SCD; of which 15.7% were fragility fractures. After adjusting for age and hypogonadism, low bone mass was associated with an increased overall fracture prevalence (OR: 1.8, 95% CI: 1.03, 3.23; p=0.041), but not with fragility fracture. In contrast, abnormal bone quality was strongly associated with fragility fracture after adjustment for age, sex, and BMI (OR: 11.4, 95% CI: 2.2, 59.1, p=0.004).

**Conclusions:** Bone quality by TBS may be a valuable tool in predicting the risk of fragility fractures in young adults with hemoglobinopathies and should be considered when making decisions for anti-resorptive therapy in those with low BMD naive to fracture or where disease-specific artifacts complicate accurate spine assessment by BMD alone.

## INTRODUCTION

Thalassemia (Thal) and Sickle Cell Disease (SCD) are the two most common genetic diseases worldwide associated with defective hemoglobin synthesis. Individuals with Thal and SCD are at high risk for low bone mass. Up to 36% of adults with SCD and 64% of adults with Thal are reported to have low bone mass (^1,2^). Though the exact mechanisms explaining the prevalence of low bone mass in these populations are not fully understood, several studies implicate erythroid marrow hyperplasia, iron deposition, chelator toxicity, hypogonadism, diabetes, vitamin D and zinc deficiencies, low muscle mass, and limited activity (^3,4,5^).

Low bone mass prevalence has been shown to increase with age in both Thal and SCD (^6,7,8^) and is related to an increased risk of fracture in Thal (^9,6^). Despite being at low risk of recreational injury due to limited activity, patients with hemoglobinopathies are at increased risk for fragility fractures which have been reported in up to 10% of patients (^8^). Additionally, vertebral abnormalities have been reported in up to 35% and 46% of Thal and SCD individuals respectively (^6,10,8^).

Having an imaging modality that accurately predicts future fracture so as to avoid significant morbidity would be extremely useful. Unfortunately, Dual Energy X-ray Absorptiometry (DXA), the gold standard clinical tool for assessing low bone mass, is only a proxy for bone strength and, therefore, does not consistently predict fracture (^11^). For patients with hemoglobinopathies, in particular, there are numerous artifacts that may interfere with accurate DXA assessment. Hepatic iron overload, platyspondyly, vertebral endplate abnormalities, and vertebral bi-concavity malformations artificially increase spine bone mineral density (BMD) in Thal and SCD, while avascular necrosis in SCD increases BMD at the proximal femur (^12,13,14,15^); each contributing to greater challenges with fracture risk assessment.

Trabecular bone score (TBS), a textural analysis of bone quality derived from lumbar spine scans by DXA, may serve as a proxy for bone microarchitecture and provide skeletal information that is not captured from standard BMD measurements (^16^). Over the past decade, numerous studies in healthy populations and in those with chronic disease have found TBS to be more predictive of fracture than BMD alone (^17, 18,19^). TBS is now recommended to be incorporated into the Fracture Risk Assessment Tool (FRAX) calculator along with BMD to determine overall fracture risk (^20^). In 2023, the International Society for Clinical Densitometry produced a set of guidelines for when to use TBS in clinical practice, though there are no recommendations for the use of this tool in populations that suffer fracture at a young age, or who have such challenges to the interpretation of DXA imaging, such as those with Thal or SCD (^20^).

Bone quality has been assessed previously in patients with Thal, and it has been found to be reflective of hypogonadism and predictive of vertebral fracture (^21,10,9^). This year, Seiller et al^8^ also explored bone quality in a group of French patients with sickle cell disease but found no association with fracture risk. The current study aims to further explore the relationship between bone density and bone quality in patients with hemoglobinopathies and to determine whether TBS alone or in combination with BMD by DXA may offer a more accurate assessment of bone quality and fracture risk.

## METHODS

A retrospective chart review was conducted for patients followed at the University of California, San Francisco (UCSF) Benioff Children’s Hospital Oakland (BCH-Oakland) hematology clinic with a diagnosis of thalassemia (β-thal, E-β thal, HbH or HbH/Constant Spring) or sickle cell disease (HbSS, HbSC or HbSβ°Thal). Patients were included if they had at least one spine bone mineral density (BMD) scan performed by DXA (Hologic, Horizon A, Bedford, MA) in the previous 10 years, were ≥ 12 years of age, and weighed >40 kg at the time of the examination. For eligible patients with more than one bone density assessment during this period, the most recent spine scan, aligned with corresponding hip BMD images, was queried from the Bone Density Clinic database and used for analysis along with patient weight, height, and estimated total calcium intake (diet + supplementation) at the time of exam. Patient report of fracture was gathered at each DXA exam, verified by medical record when possible, and included in the clinic database. Each spine scan was re-analyzed using the trabecular bone score software program (TBS iNsight, Medimaps SASU, Mérignac, France), and TBS for L1-L4 was recorded. According to ISCD recommendations, any lumbar scan that did not include all 4 vertebrae was excluded from this analysis (ISCD 2023 Official Positions). Information regarding patient diagnosis, gender, age, hypogonadism, hypothyroidism or diabetes diagnoses, transfusion dependence, liver iron concentration, and 25OH vitamin D level were abstracted from the medical record. A convenience sample of healthy controls who had participated in prior non-interventional bone health research studies at BCH-Oakland was also included in this analysis (^22, 23^). Similar data were available from the control subjects except for iron assessments. This study was reviewed and approved by the Investigative Review Board at the UCSF Benioff Children’s Hospital Oakland (IRB#2016-050).

Information pertinent to a fracture (age of patient, skeletal location, cause) was abstracted from the clinic database and categorized. Site of skeletal fracture was categorized into 5 broad categories: ‘Upper’ extremity fractures (humerus, radius, ulna, elbow, hand), ‘Lower’ extremity fractures (femur, tibia, fibula, patella, foot and pelvis), ‘Spine’ (any vertebral fracture), fractures that occurred primarily along with axial skeleton noted as ‘SCP’’ (skull, jaw, clavicle, and ribs) and Other/Unknown. Fractures of fingers and toes were excluded from the analysis. Etiology of fracture was also categorized into 4 broad categories: *Fall:* from greater than standing height; *Recreational:* occurring during any recreational activity including bicycling ; *Fragility:* low impact fractures and those that occur from standing height or less; *MVA*: fractures sustained from motor-vehicle accidents and *Other/Unknown*: Heavy object/assault/unknown. Details of treatment were was not available for all fractures and, therefore, were not included in the analysis. Low bone mass was defined as a BMD Z-score at any site ≤-2.0. Bone quality was designated as optimal (TBS ≥ 1.35), subnormal (TBS 1.34 - 1.20), or abnormal (TBS <1.20). Given the lack of race specific reference data available for TBS, raw data and cut-off values were used to categorize bone quality rather than conversion to TBS Z-scores. Recent reports confirm the validity of TBS for fracture risk stratification in ethnically diverse populations (^24^). Spine scans were excluded if they contained interfering artifacts (e.g. gallstones, renal stones, belly button rings, gastrostomy tubes).

### Data Analysis

For the purposes of this report, we recorded vitamin D deficiency (Total 25OH vitamin D < 20 ng/mL), hypogonadism (prescribed estrogen or androgen replacement therapy), hypothyroidism (prescribed thyroid hormone replacement therapy), diabetes (prescribed insulin or oral hypoglycemic agents), and transfusion dependency (≥8 transfusions per year). Adequacy of treatment for these conditions was not addressed though all subjects were under routine medical care. Differences in continuous variables between groups (Thal, SCD, Controls) were analyzed by ANOVA and Student’s t-tests for unpaired comparisons. For categorical data (race, sex at birth, disease type, fracture site, and cause), differences between groups were analyzed using Pearson’s chi-squared test, with Fisher’s exact test for comparisons of small samples. The main outcomes of interest were fracture prevalence by group, etiology of fracture by group, and factors related to fracture prevalence and etiology. Correlations between TBS, BMD, and age were explored using Pearson’s correlation coefficient. Multivariate logistic regression analyses were used to evaluate the effects of age, gender, hematologic parameters, endocrinopathies (hypogonadism, hypothyroidism), BMD, and TBS on fracture prevalence and then, more specifically, on etiology of fracture. A p-value of <0.05 was deemed significant for all tests, with p<0.10 considered a trend.

## RESULTS

Data from 360 individuals were included in this retrospective review, including 126 patients with Thal, 170 with SCD and 64 healthy controls (**Table 1**). Reflective of our patient population at BCH-Oakland, the majority of the patients with Thal had a diagnosis of ꞵ-thalassemia followed by HbE-ꞵ thalassemia, while the majority of patients with SCD were homozygous SS. In this sample, patients with Thal were significantly older than SCD or Controls, while fewer males were included in the control group. As expected, the majority of the Thal patients were Caucasian and Asian, while 91% of the patients with SCD were African American. More patients with Thal were chronically transfused, though the hepatic iron burden was significantly higher in SCD compared to Thal (p=0.036). Patients with SCD had significantly lower 25OH vitamin D levels compared to Thal and controls (p<0.01 for each comparison). The prevalence of endocrinopathies (hypogonadism 31.4%, hypothyroidism 17.7% and diabetes 12.5%) was almost exclusive to the Thal group.

**Table 1.** Subject Demographics and Clinical Characteristics by Diagnosis.

| Characteristics | Thalassemia<br>n=126 | Sickle Cell<br>Disease<br>n=170 | Healthy Controls<br>n=64 | p-value |
| --- | --- | --- | --- | --- |
| Age, years | 31.7 ± 11.9<br>(12.0 , 63.1) | 24.6 ± 13.5<br>(12.0 , 69.7) | 25.9 ± 8.0<br>(12.1, 44.0) | <0.001 |
| Sex at Birth<br>Male/Female, % | 51.6%, 48.4% | 42.9%, 57.1% | 17.2%, 82.8% | <0.001 |
| Race, %<br>Caucasian<br>African American<br>Asian<br>Hispanic<br>Mixed/Other | 10.0%<br>-<br>70.0%<br>0.8%<br>19.2% | -<br>91.0%<br>-<br>4.2%<br>4.8% | 45.3%<br>14.1%<br>29.7%<br>3.1%<br>7.8% | <0.001 |
| Diagnosis, % | β-Thal Major 53.3%<br>β-Thal Intermedia 9.8%<br>Eβ-Thal 23.0%<br>HbH or HbH/CS 13.9% | Hb-SS 77.4%<br>Hb-SC 13.7%<br>Sβ<sup>o</sup>-Thal 8.9% | N/A |  |
| Transfusion<br>Dependent, % | 73.3% | 31.8% | N/A | <0.001 |
| LIC, ug Fe/g wet tissue | 1882 ± 1862<br>(110, 7096) | 2676 ± 2134<br>(81, 7829) | N/A | 0.036 |
| Hypogonadism, % | 31.4% | <1% | 0% | <0.001 |
| Hypothyroidism, % | 17.7% | 0% | 0% | <0.001 |
| Diabetes, % | 12.5% | 3.3% | 0% | <0.001 |
| BMI, kg/m<sup>2</sup> | 22.5 ± 3.8<br>(16.2, 41.2) | 23.0 ± 4.4<br>(14.7, 38.0) | 24.1 ± 4.9<br>(17.2, 40.4) | 0.058 |
| BMI > 30 kg/m<sup>2</sup> | 4.0% | 6.5% | 12.9% | 0.081 |
| Calcium intake, mg/day | 596 ± 468<br>(100 , 2200) | 616 ± 449<br>(100 , 2200) | 664 ± 358<br>(159 , 1501) | NS |
| 25OH Vitamin D, ng/mL | 32.4 ± 13.6<br>(11.6 , 70.6) | 24.2 ± 8.6<br>(7.0 , 48.8) | 27.6 ± 8.8<br>(9.0 , 54.0) | <0.001 |
| 25OHD Deficiency,<br>< 20 ng/mL, % | 18.9% | 29.5% | 20.7% | NS |
| Spine BMD Z-score | -2.2 ± 1.1<br>(-5.7, 1.3) | -1.4 ± 1.5<br>(-4.9, 4.8) | -0.2 ± 0.8<br>(-1.9, 1.7) | <0.001 |
| Hip BMD Z-score | -1.6 ± 1.0<br>(-4.8, 1.0) | -0.7 ± 1.1<br>(-0.7, 1.1) | -0.1 ± 0.9<br>(-2.0, 2.3) | <0.001 |
| Low bone mass, % | 62.7% | 35.9% | 3.1% | <0.001 |
| Trabecular bone score | 1.28 ± 0.12<br>(0.86, 1.61) | 1.33 ± 0.11<br>(1.02, 1.74) | 1.46 ± 0.08<br>(1.26, 1.63) | <0.001 |
| TBS Category |  |  |  |  |
| Optimal, % | 30.1% | 41.1% | 95.1% | <0.001 |
| Subnormal, % | 43.9% | 51.8% | 4.9% |  |
| Abnormal, % | 26.0% | 7.1% | 0% |  |

Continuous variables presented as Mean±SD (range)
Categorical variables presented as percentages of total sample for each diagnosis
P-value for continuous variables is from ANOVA, for categorical by chi-squared with fisher exact
BMI: body mass index; BMD: Bone mineral density
HbH or HbH/CS: hemoglobin H thalassemia/constant spring
LIC: Liver iron concentration assessed by SQUID
TBS: Trabecular bone score

As reported previously, the spine BMD Z-score was consistently lower than the hip BMD Z-score in both Thal and SCD groups (Table 1)(^3,8^). Significantly more patients with Thal (63%) had low bone mass [defined as BMD Z-score <-2.0 at either the hip or spine] compared to SCD (36%) or controls (3%, p<0.001). Removing controls from the analysis, bone quality was positively correlated with spine BMD (r=0.58, p<0.001), while low spine BMD Z-score was also strongly associated with poor bone quality in hemoglobinopathies (Chi squared, p<0.001). Subjects with low bone mass were more likely to be hypogonadal, transfusion-dependent, and have a lower BMI (all p<0.01) irrespective of age, gender, or vitamin D level. Abnormal bone quality (defined as TBS <1.2) was also observed in a higher percentage of patients with Thal (26.0%) compared to SCD (7.1%, p=0.001). Subjects with poor bone quality were more likely to be older, hypogonadal, diabetic, and transfusion-dependent with higher vitamin D levels (all p<0.001).

Overall, 25.7% of patients with hemoglobinopathies (76 out of 296) had sustained a fracture in their lifetime (**Table 2**). Fracture prevalence was more common in Thal (35.6%) compared to controls (16.4%) but not different from SCD (22.9%). For the group as a whole, there was an overall difference in fracture prevalence by gender (p=0.001, ANOVA). Within the SCD group, fracture was more common in females compared to males (p=0.015). For those who had fractured, there were no significant differences in the number of fractures or the age at first fracture between groups. There were also no significant differences in the location of fractures by group, though interestingly, the healthy controls only sustained upper or lower extremity fractures. There were significant differences noted in the cause of fracture by group (p=0.047). The majority (55%) of the fractures sustained in healthy controls were while playing sports or participating in various physical activities, while 21% of all fractures in patients with Thal and 15% of fractures in SCD were categorized as low impact/fragility type fractures. None of the controls sustained fragility fractures.

**Table 2:** Summary of All Fracture Characteristics by Diagnosis Group.

|  | Thalassemia<br>n=135 | Sickle Cell Disease<br>n=175 | Healthy Controls<br>n=67 | p-value |
| --- | --- | --- | --- | --- |
| Fracture Prevalence,<br>All subjects, % | 35.6% | 22.9% | 16.4% | 0.005 |
| <sup>a</sup> Females | 45.8% | 75.0% | 36.4% | 0.007 |
| Males | 54.2% | 25.0% | 63.6% |  |
| <sup>b</sup> Multiple Fracture, % | 7.1% | 1.0% | 4.7% | NS |
| Total Number of Fractures | 48 | 40 | 11 |  |
| <sup>c</sup> Average Number of<br>Fractures | 1.2 ± 0.6<br>(0 , 3) | 1.0 ± 0.6<br>(0 , 3) | 1.2 ± 0.7<br>(0 , 2) | NS |
| Age at First Fracture,<br>years | 20.0 ± 14.9<br>(2 , 56) | 19.1 ± 14.9<br>(1 , 55) | 10.6 ± 4.0<br>(4 , 15) | NS |
| Location of Fracture |  |  |  | NS |
| Upper Extremity | 36% | 35% | 45% |  |
| Lower Extremity | 45% | 35% | 55% |  |
| Spine | 2% | 3% | 0% |  |
| Skull, Clavicle, Ribs | 15% | 18% | 0% |  |
| Other/Unknown | 2% | 10% | 0% |  |
| Cause of Fracture |  |  |  | 0.047 |
| Fall | 19% | 28% | 45% |  |
| Fragility | 21% | 15% | 0% |  |
| MVA | 11% | 20% | 0% |  |
| Recreational | 23% | 25% | 55% |  |
| Other/Unknown | 26% | 13% | 0% |  |

Continuous variables presented as Mean ± SD (range)
Categorical variables presented as a percentage of total sample for each diagnosis
P-value for continuous variables by ANOVA, for categorical by chi-squared with fisher exact
<sup>a</sup> Percentage of those who fractured who were female/male by diagnostic group
<sup>b</sup> Percentage of subjects who sustained more than 1 fracture
<sup>c</sup> Average number of fractures and age at first fracture, each subject only captured once

Focusing the analyses on Thal and SCD subjects only, those who had sustained a fracture were more likely to be older and transfusion-dependent (**Table 3**). Those with fracture history also had higher vitamin D levels but lower hip BMD Z-scores with a tendency for low spine BMD Z-scores, poor bone quality, and hypogonadism. Using multivariate logistic regression, after adjusting for age and hypogonadism, low BMD was associated with increased fracture prevalence (OR: 1.8, 95% CI: 1.03, 3.23; p=0.041). Only a trend was observed between poor bone quality and overall fracture prevalence (p=0.10).

**Table 3:**
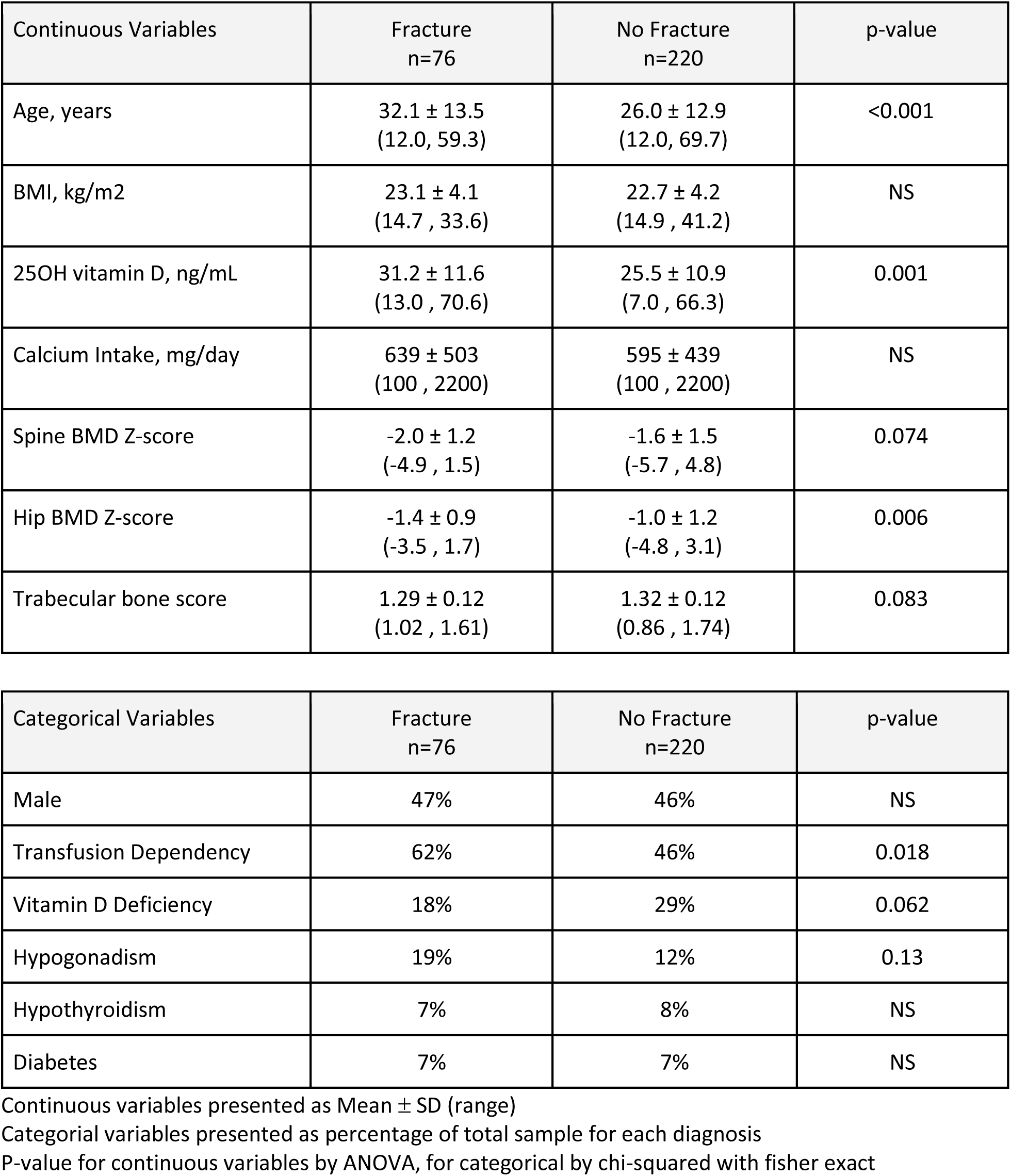
Factors Related to Fracture for Individuals with Thalassemia or Sickle Cell Disease (n=296), Unadjusted Relationships.

Additional analyses were then made exploring factors associated with fracture type. Low bone mass was not related to the cause of fracture either in the group as a whole or in the select group of patients with hemoglobinopathies. In contrast, low TBS was associated with the cause of fracture in the combined group of Thal and SCD patients, p<0.001 (**Figures 1a,b**). The youngest age for sustaining a fragility fracture was 8 years in a subject with HbSS (vertebral compression fracture). The oldest patient was an adult female with transfusion dependent B-thalassemia who sustained 3 fragility fractures between 56 to 58 years of age in her upper and lower extremities. All fractures in the Thal and SCD groups were then collapsed into 2 categories: fragility (n=12) and non-fragility fractures (n=64). So as to not bias the results, subjects were counted in this analysis only once based on their most significant fracture. Subjects with fragility fractures were more likely to be female, older, with a higher BMI, and poor bone quality (**Table 4**). Using multivariate analysis, abnormal bone quality was associated with 11-fold greater odds of having a fragility fracture in patients with hemoglobinopathies after adjusting for age, gender, and BMI (OR: 11.4, 95% CI: 2.2, 59.1, p=0.004). Patient diagnosis (Thal/SCD), age, vitamin D level, hypogonadism, hypothyroidism, and transfusion dependency were not significantly associated with fragility fracture in these multivariate models. Interestingly, low bone mass was not associated with fragility fracture in these subjects, even after adjustment for age or gender.

**Figure 1a:**
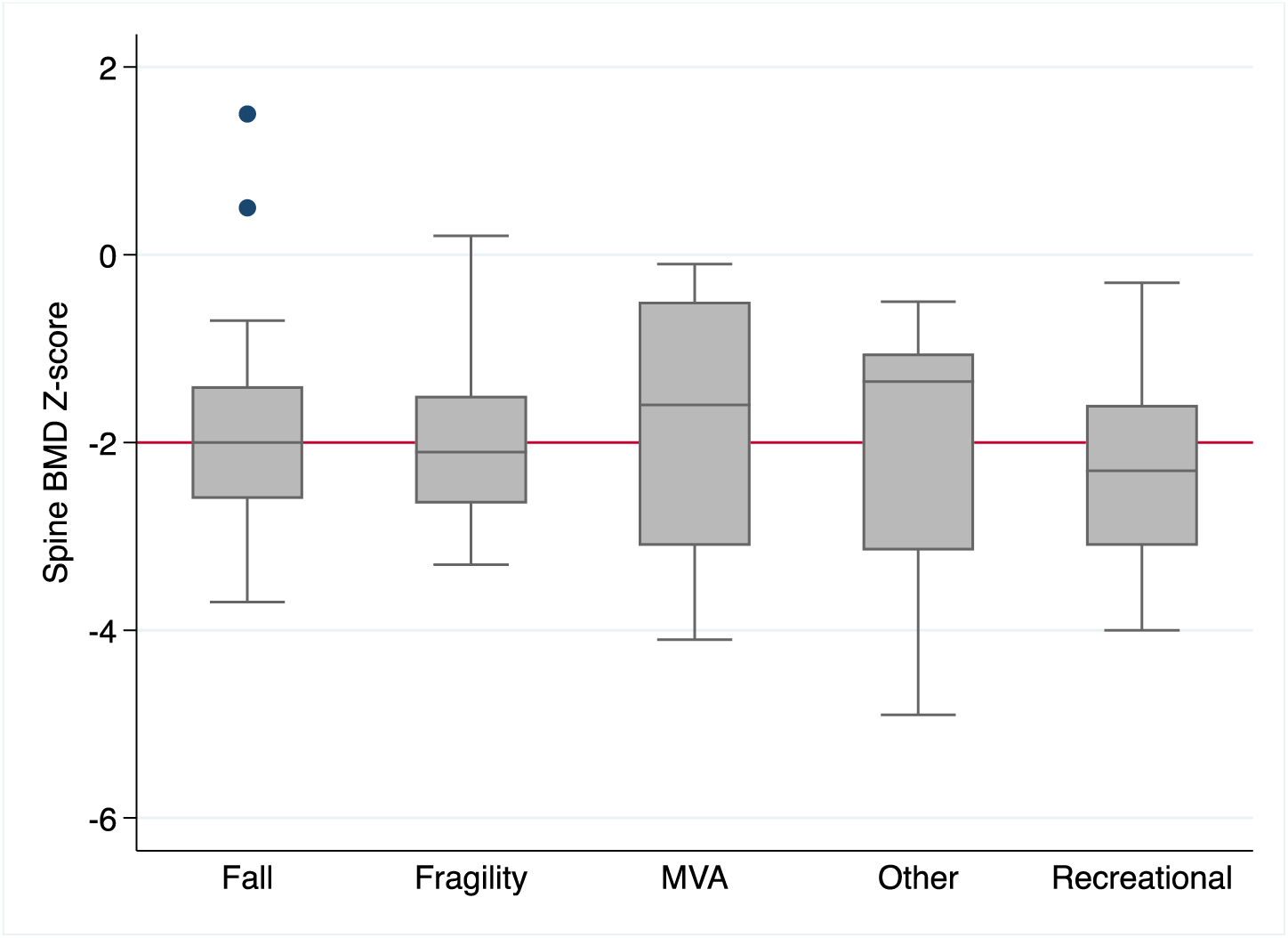
Spine BMD Z-score by Cause of Fracture for Thal and SCD Subjects who Have Sustained a Fracture (n=76) (Below Reference Line=low bone mass), p=NS

**Figure 1b:**
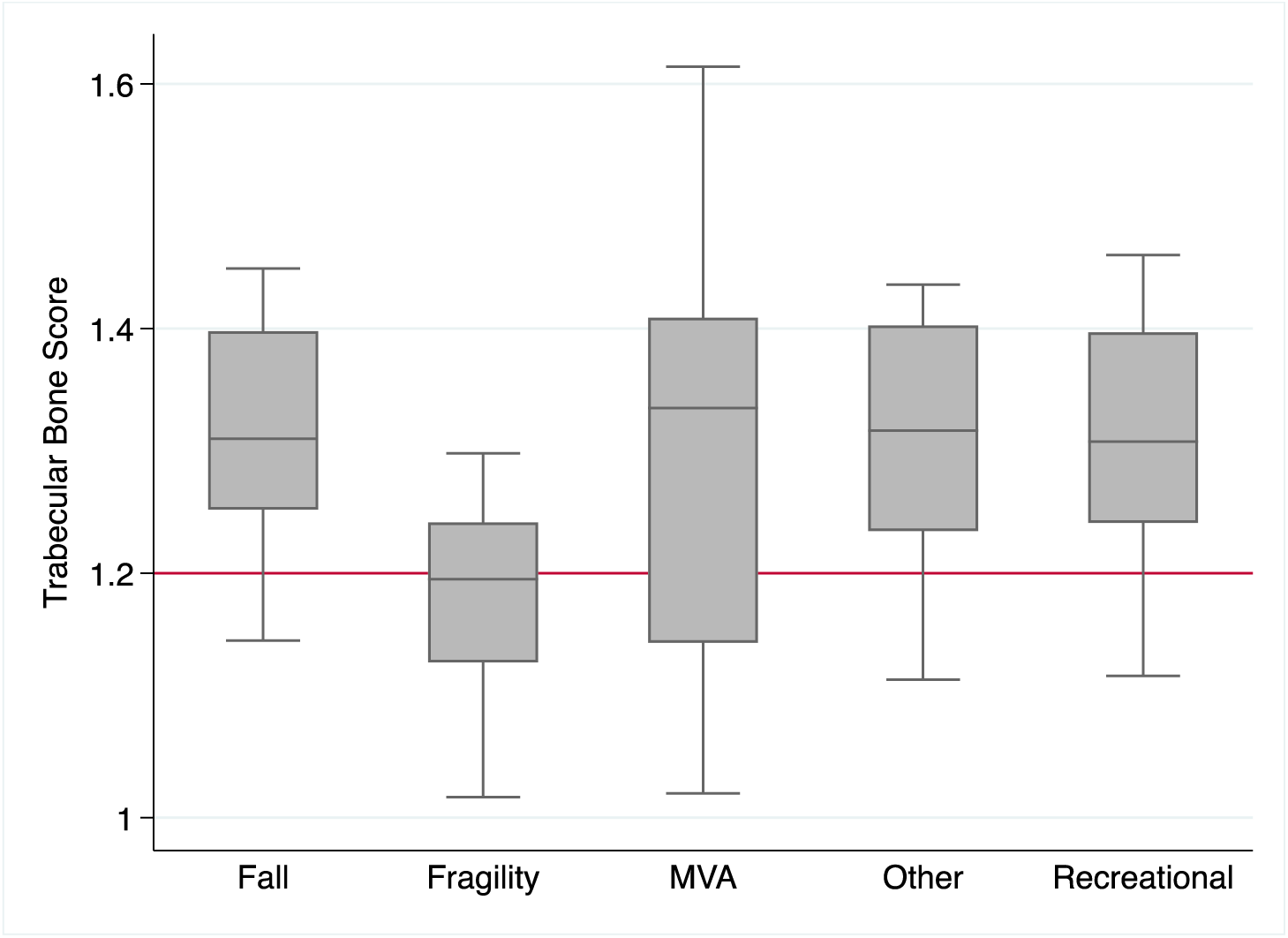
Trabecular Bone Score by Cause of Fracture, for Thal and SCD Subjects who Have Sustained a Fracture (n=76) (Below Reference Line=abnormal bone quality), p<0.001 p-value from chi squared association between low TBS and Cause Fracture

**Table 4:** Factors Related to Fragility Fracture in Individuals with a History of Fracture Unadjusted relationships for Thal and SCD Subjects Only (n=76)

| Continuous Variables | Fragility Fracture<br>n=12 | Other Fracture<br>n=64 | p-value |
| --- | --- | --- | --- |
| Age at time of fracture, years | 32.2 ± 19.7<br>(8.0 , 58.0) | 17.4 ± 12.2<br>(1.0 , 55.0) | <0.001 |
| Age at time of DXA, years | 41.6 ± 15.2<br>(17.3 , 59.0) | 30.0 ± 12.4<br>(12.0 , 59.3) | 0.005 |
| BMI, kg/m<sup>2</sup> | 25.7 ± 3.5<br>(20.5 , 33.3) | 22.5 ± 4.0<br>(14.7 , 33.6) | 0.014 |
| 25OH vitamin D, ng/mL | 33.4 ± 10.3<br>(13.0 , 41.9) | 30.8 ± 11.9<br>(13.0 , 70.6) | NS |
| Calcium Intake, mg/day | 697 ± 584<br>(100 , 2150) | 633 ± 493<br>(100 , 2200) | NS |
| Spine BMD Z-score | -2.0 ± 0.9<br>(-3.3 , 0.2) | -2.0 ± 1.2<br>(-4.9 , 1.5) | NS |
| Hip BMD Z-score | -1.6 ± 0.7<br>(-3.2 , -0.4) | -1.4 ± 0.9<br>(-3.5 , 1.7) | NS |
| Trabecular bone score | 1.18 ± 0.08<br>(1.02 , 1.30) | 1.31 ± 0.11<br>(1.02 , 1.61) | 0.002 |

| Categorical Variables | Fragility Fracture<br>n=12 | Other Fracture<br>n=64 | p-value |
| --- | --- | --- | --- |
| Male | 17% | 52% | 0.023 |
| Transfusion Dependency | 67% | 62% | NS |
| Vitamin D Deficiency | 18% | 17% | NS |
| Hypogonadism | 25% | 18% | NS |
| Hypothyroidism | 9% | 7% | NS |
| Diabetes | 9% | 0% | NS |

Continuous variables presented as Mean ± SD
Categorical variables presented as a percentage of the total sample for the fracture and non-fracture groups
P-value for continuous variables by ANOVA, for categorical by chi-squared with fisher exact

## DISCUSSION

This is the first study to assess both bone quantity and quality in a combined group of patients with hemoglobinopathies and explore their relationship to overall fracture prevalence. Nearly half of the patients with hemoglobinopathies in this cohort were categorized with low bone mass while only 15% had abnormal bone quality. Though low bone mass was weakly associated with overall fracture prevalence, only low TBS was significantly associated with fragility fracture. This discrepancy suggests that bone quality plays a greater role in determining fragility fracture risk than bone quantity in these patients. As optimal fracture risk assessment is dependent on the ability to accurately assess true bone strength, identifying clinical tools that better assess the microarchitecture of bone, such as trabecular bone score, could provide more information on when to recommend intervention. We propose incorporating TBS into the bone health assessment for a more robust assessment of fracture risk in hemoglobinopathies.

Data from this cohort suggests that despite low levels of reported recreational activity, fracture is common in patients with hemoglobinopathies with a prevalence rate of 25%. Our ability to anticipate who may be at greatest risk of fracture remains a challenge. There are multiple difficulties in bone health assessment by DXA in patients with hemoglobinopathies. We have previously shown that in patients with hepatic iron overload, BMD at L1 and L2 may be artificially elevated by iron deposition in the liver (^12^). Others have detailed how biconcavity vertebral height abnormalities, present in up to 35% of Thal and 46% of SCD may lead to artificially increased BMD, while end-plate abnormalities and platyspondyly are common, though not necessarily associated with poor bone quality (^10,8,15^). Furthermore, avascular osteonecrosis, a common morbidity found in the proximal femur of up to 22% of SCD is associated with increased BMD by DXA (^14,25^). Therefore, it may be difficult to assess ‘true’ bone strength by BMD assessment alone.

Fracture prevalence in this cohort of Thal (35.6%) and SCD (22.9%) subjects was similar to that previously published by our group and others where patients with Thal have reports of between 36 to 38% lifetime fracture risk, while patients with SCD have reports of 20 to 30% (^14,26,27^). In our prior study, subjects were pre-selected for extremely high iron levels, which likely contributed to bone morbidity (^26^). However, in the current cohort, only a third of SCD subjects were chronically transfused, and hepatic iron burden in Thal was moderate for most, suggesting other etiologies for increased fracture risk in this population, such as disease specific morbidity, nutritional deficiencies, and low physical activity.

Given this cohort of patients with hemoglobinopathies was relatively young (average age 27 years), we would expect few fractures to have occurred from non-traumatic, non-sports related injuries, yet 21% of all Thal fractures and 15% of all SCD fractures were categorized as low impact/fragility type. A recent report on French patients with SCD found a similar rate of fragility fracture in 138 adult patients (10%), but they found no relationship of low bone mass or bone quality to fracture risk (^8^). Perhaps their smaller sample size, or monocentric sample of subjects with SCD, could account for their inability to relate BMD or TBS to fragility fracture. Thavonlun et al (^28^) observed a similar rate of fragility fracture in transfusion dependent Thal (20%), and observed a strong association with sarcopenia. Though we did not measure sarcopenia directly, BMI was significantly higher in those who sustained a fragility fracture suggesting body composition may play a mechanistic role. Though others have found bone quality to be predictive of vertebral fracture (^10,9^), to our knowledge this is the first study to correlate TBS to low impact fragility fracture in hemoglobinopathies.

Fracture risk is related to both modifiable (lifestyle factors, medications, disease characteristics, balance, bone density, bone quality) and non-modifiable factors (age, sex, ethnicity, genetics). A number of studies have shown that lifestyle and disease characteristics can be modified to improve bone density and thereby reduce the possibility of fracture in hemoglobinopathies. Bisphosphonates, denosumab, zinc, strontium, hypercaloric products and vibration therapy are just a few examples of therapies that have been shown or are currently being tested to increase BMD in Thal or SCD ( ^29,30,31,32,33^). To our knowledge, there have been no clinical trials focused specifically on improving bone quality in hemoglobinopathies. New software tools using DXA instrumentation have recently become available that assess not only bone quantity and quality but also bone strength and fatigue (Bone Strength Index,^34^). Future research focused on how modifying lifestyle factors in patients with hemoglobinopathies may not only improve bone quantity but also enhance bone quality and possibly bone strength is needed.

As this was a retrospective sample of patients from our clinic, there are limitations. All adolescent and adult patients who had a DXA exam within the prior decade were included, therefore we had no control over the patient characteristics. Subjects with Thal were older than SCD and Controls, and as age is related to fracture, where feasible, our multivariate analyses were adjusted for age. Vitamin D supplementation is routinely prescribed to patients in our clinic with low bone mass. Therefore, it was not surprising to observe a relationship between high vitamin D and low BMD. As all fractures occurred prior to the imaging assessments, we were unable to draw temporal relationships between the fracture and the bone density or bone quality measurements. Given the rarity of fracture as an outcome, this type of study design is not unique (^26,35,28^), though we recognize only associations and not causation between fracture and bone health can be made with these data. Finally, though the data set includes a relatively large number of subjects for this population of patients, the total number of fractures, particularly fragility fractures, is small, limiting the number of covariate analyses. What is unique to this study is the assessment of both patients with Thal and SCD who are at increased risk of fracture, combined with a diverse population of healthy controls for comparison.

## CONCLUSIONS

This study supports a relationship between reduced bone mass and bone quality in adolescent and adult patients with hemoglobinopathies. Though reduced bone mass was associated with overall fracture prevalence it could not be used to differentiate those with fragility fracture. In contrast, poor bone quality was strongly associated with a history of fragility fracture. While DXA remains the primary tool for assessing low bone mass in hemoglobinopathies, these data suggest its limitations for characterizing bone strength. Bone quality by TBS may be valuable as part of bone health assessments in adults with hemoglobinopathies and should be considered in decisions regarding anti-resorptive therapy in those with very low BMD who are naive to fracture. Our study indicates that particular attention should be paid to strategies that preserve or enhance bone quality to avoid fragility fractures.

## Data Availability

All data produced in the present study are available upon reasonable request to the authors

## Acknowledgements

We would like to thank the students who contributed to the data collection as part of the Summer Student Research Program (SSRP), Ms. Melissa Cervantes and Ms. Kaylianna Cadena.

## Citations

1. Ananvutisombat N, Tantiworawit A, Punnachet T, et al. Prevalence and risk factors predisposing low bone mineral density in patients with thalassemia. Front Endocrinol. 2024;15:1393865. doi:10.3389/fendo.2024.1393865

2. Garadah TS, Hassan AB, Jaradat AA, et al. Predictors of Abnormal Bone Mass Density in Adult Patients with Homozygous Sickle-Cell Disease. Clin Med Insights Endocrinol Diabetes. 2015;8:CMED.S24501. doi:10.4137/CMED.S24501

3. Wong P, Fuller PJ, Gillespie MT, Milat F. Bone Disease in Thalassemia: A Molecular and Clinical Overview. Endocrine Reviews. 2016;37(4):320–346. doi:10.1210/er.2015-1105

4. Di Paola A, Marrapodi MM, Di Martino M, et al. Bone Health Impairment in Patients with Hemoglobinopathies: From Biological Bases to New Possible Therapeutic Strategies. IJMS. 2024;25(5):2902. doi:10.3390/ijms25052902

5. Giordano P, Urbano F, Lassandro G, Faienza MF. Mechanisms of Bone Impairment in Sickle Bone Disease. IJERPH. 2021;18(4):1832. doi:10.3390/ijerph18041832

6. Vogiatzi MG, Macklin EA, Fung EB, et al. Bone Disease in Thalassemia: A Frequent and Still Unresolved Problem. Journal of Bone and Mineral Research. 2009;24(3):543–557. doi:10.1359/jbmr.080505

7. Adesina OO, Gurney JG, Kang GL, et al. Height-corrected low bone density associates with severe outcomes in sickle cell disease: SCCRIP cohort study results. Blood Advances. 2019;3(9):1476–1488. doi:10.1182/bloodadvances.2018026047

8. Seiller J, Merle B, Fort R, et al. Prevalence of bone complications in young patients with sickle cell disease presenting low bone mineral density. Bone. 2024;178:116924. doi:10.1016/j.bone.2023.116924

9. Teawtrakul N, Chukanhom S, Charoensri S, Somboonporn C, Pongchaiyakul C. The Trabecular Bone Score as a Predictor for Thalassemia-Induced Vertebral Fractures in Northeastern Thailand. Anemia. 2020;2020:1–5. doi:10.1155/2020/4634709

10. Osella G, Priola AM, Priola SM, et al. Dual-Energy X-ray Absorptiometry Predictors of Vertebral Deformities in Beta-Thalassemia Major. Journal of Clinical Densitometry. 2018;21(4):507–516. doi:10.1016/j.jocd.2017.06.028

11. McCloskey E, Tan ATH, Schini M. Update on fracture risk assessment in osteoporosis. *Current Opinion in Endocrinology*, Diabetes & Obesity. Published online May 30, 2024. doi:10.1097/MED.0000000000000871

12. Allard HM, Calvelli L, Weyhmiller MG, Gildengorin G, Fung EB. Vertebral Bone Density Measurements by DXA are Influenced by Hepatic Iron Overload in Patients with Hemoglobinopathies. Journal of Clinical Densitometry. 2019;22(3):329–337. doi:10.1016/j.jocd.2018.07.001

13. Serarslan Y, Kalacı A, Özkan C, Doğramacı Y, Çokluk C, Yanat AN. Morphometry of the thoracolumbar vertebrae in sickle cell disease. Journal of Clinical Neuroscience. 2010;17(2):182–186. doi:10.1016/j.jocn.2009.05.010

14. De Luna G, Ranque B, Courbebaisse M, et al. High bone mineral density in sickle cell disease: Prevalence and characteristics. Bone. 2018;110:199–203. doi:10.1016/j.bone.2018.02.003

15. Pellegrino F, Zatelli MC, Bondanelli M, et al. Dual-energy X-ray absorptiometry pitfalls in Thalassemia Major. Endocrine. 2019;65(3):469–482. doi:10.1007/s12020-019-02003-x

16. Shevroja E, Lamy O, Kohlmeier L, Koromani F, Rivadeneira F, Hans D. Use of Trabecular Bone Score (TBS) as a Complementary Approach to Dual-energy X-ray Absorptiometry (DXA) for Fracture Risk Assessment in Clinical Practice. Journal of Clinical Densitometry. 2017;20(3):334–345. doi:10.1016/j.jocd.2017.06.019

17. Borgen TT, Bjørnerem Å, Solberg LB, et al. High prevalence of vertebral fractures and low trabecular bone score in patients with fragility fractures: A cross-sectional sub-study of NoFRACT. Bone. 2019;122:14–21. doi:10.1016/j.bone.2019.02.008

18. Leslie WD, Binkley N, Hans D. Effects of Lumbar Spine Vertebral Fractures on Trabecular Bone Score (TBS): The Manitoba BMD Registry. Journal of Clinical Densitometry. 2024;27(4):101533. doi:10.1016/j.jocd.2024.101533

19. Shevroja E, Reginster JY, Lamy O, et al. Update on the clinical use of trabecular bone score (TBS) in the management of osteoporosis: results of an expert group meeting organized by the European Society for Clinical and Economic Aspects of Osteoporosis, Osteoarthritis and Musculoskeletal Diseases (ESCEO), and the International Osteoporosis Foundation (IOF) under the auspices of WHO Collaborating Center for Epidemiology of Musculoskeletal Health and Aging. Osteoporos Int. 2023;34(9):1501–1529. doi:10.1007/s00198-023-06817-4

20. Goel H, Binkley N, Boggild M, et al. Clinical Use of Trabecular Bone Score: The 2023 ISCD Official Positions. Journal of Clinical Densitometry. 2024;27(1):101452. doi:10.1016/j.jocd.2023.101452

21. Baldini M, Marcon A, Ulivieri FM, et al. Bone quality in beta-thalassemia intermedia: relationships with bone quantity and endocrine and hematologic variables. Ann Hematol. 2017;96(6):995–1003. doi:10.1007/s00277-017-2959-0

22. Fung EB, Vichinsky EP, Kwiatkowski JL, et al. Characterization of low bone mass in young patients with thalassemia by DXA, pQCT and markers of bone turnover. Bone. 2011;48(6):1305–1312. doi:10.1016/j.bone.2011.03.765

23. Cullers A, King JC, Van Loan M, Gildengorin G, Fung EB. Effect of prenatal calcium supplementation on bone during pregnancy and 1 y postpartum. The American Journal of Clinical Nutrition. 2019;109(1):197–206. doi:10.1093/ajcn/nqy233

24. Leslie WD, Binkley N, Hans D. Ethnicity and Fracture Risk Stratification from Trabecular Bone Score in Canadian Women: The Manitoba BMD Registry. Journal of Clinical Densitometry. 2023;26(1):83–89. doi:10.1016/j.jocd.2022.12.004

25. Adesina O, Brunson A, Keegan THM, Wun T. Osteonecrosis of the femoral head in sickle cell disease: prevalence, comorbidities, and surgical outcomes in California. Blood Advances. 2017;1(16):1287–1295. doi:10.1182/bloodadvances.2017005256

26. Fung EB, Harmatz PR, Milet M, et al. Fracture prevalence and relationship to endocrinopathy in iron overloaded patients with sickle cell disease and thalassemia. Bone. 2008;43(1):162–168. doi:10.1016/j.bone.2008.03.003

27. Arlet JB, Courbebaisse M, Chatellier G, et al. Relationship between vitamin D deficiency and bone fragility in sickle cell disease: A cohort study of 56 adults. Bone. 2013;52(1):206–211. doi:10.1016/j.bone.2012.10.005

28. Thavonlun S, Houngngam N, Kingpetch K, et al. Association of osteoporosis and sarcopenia with fracture risk in transfusion-dependent thalassemia. Sci Rep. 2023;13(1):16413. doi:10.1038/s41598-023-43633-6

29. Fung EB, Gariepy CA, Sawyer AJ, Higa A, Vichinsky EP. The effect of whole body vibration therapy on bone density in patients with thalassemia: a pilot study. Am J Hematol. 2012;87(10):E76–79. doi:10.1002/ajh.23305

30. Fung EB, Kwiatkowski JL, Huang JN, Gildengorin G, King JC, Vichinsky EP. Zinc supplementation improves bone density in patients with thalassemia: a double-blind, randomized, placebo-controlled trial. American Journal of Clinical Nutrition. 2013;98(4):960–971. doi:10.3945/ajcn.112.049221

31. Condé M, Lespessailles E, Wanneveich M, Allemandou D, Boulain T, Dimitrov G. Effect of nutritional supplementation on bone mineral density in children with sickle cell disease: protocol for an open-label, randomised controlled clinical trial. BMJ Open. 2024;14(4):e080235. doi:10.1136/bmjopen-2023-080235

32. Bhardwaj A, Swe KM, Sinha NK, Osunkwo I. Treatment for osteoporosis in people with ß-thalassaemia. Cochrane Database Syst Rev. 2016;3:Cd010429. doi:10.1002/14651858.CD010429.pub2

33. Adesina OO, Jenkins IC, Galvão F, et al. Alendronate preserves bone mineral density in adults with sickle cell disease and osteoporosis. Osteoporos Int. Published online October 22, 2024. doi:10.1007/s00198-024-07268-1

34. Pizza IC, Bongiorno A, Pedullà M, Albano D, Sconfienza LM, Messina C. DXA: New Concepts and Tools Beyond Bone Mineral Density. Semin Musculoskelet Radiol. 2024;28(05):528–538. doi:10.1055/s-0044-1788579

35. De Franceschi L, Gabbiani D, Giusti A, et al. Development of Algorithm for Clinical Management of Sickle Cell Bone Disease: Evidence for a Role of Vertebral Fractures in Patient Follow-up. Journal of Clinical Medicine. 2020;9(5). doi:10.3390/jcm9051601

